# Patterns of Long-term Ambulatory Care Utilization Following Exposure to Physical and Non-physical Intimate Partner Violence: A Cohort Study

**DOI:** 10.1101/2025.11.06.25339672

**Authors:** Gabriel John Dusing, Chungah Kim, Beverley M. Essue, Molly Szczech, Patricia O’Campo, Nicholas Metheny

**Affiliations:** York University; Chosun University; University of Toronto; Emory University; Unity Health Toronto

## Abstract

Intimate partner violence (IPV) is a significant determinant of women’s health that can shape care needs and care-seeking over time. Evidence on long-term patterns of ambulatory care use among survivors, however, remains limited. This study examines whether exposure to physical and non-physical IPV is associated with long-term ambulatory care utilization among women in Toronto, Canada.

Data come from the 2009-2011 Neighbourhood Effects on Health and Well-being (NEHW) study linked to the National Ambulatory Care Reporting System (NACRS) through 2020. IPV exposure was categorized as none, non-physical IPV only, or both physical and non-physical IPV. Negative binomial regression models estimated incidence rates per 100,000 person-years, incorporating follow-up time as an offset and adjusting for sociodemographic and health characteristics. Among participants with at least one ambulatory care visit, we also examined urgency of presentation (Canadian Triage Acuity Scale [CTAS]) and mode of arrival.

Women exposed to both physical and non-physical IPV had higher rates of ambulatory care use compared with women reporting no IPV or non-physical IPV alone. For non-urgent or less urgent presentations (CTAS 4-5), incidence rates were 47.94 (95%CI: 32.75-63.13) per 100,000 person-years among women exposed to both forms of IPV, compared with 23.59 (95%CI: 17.34-29.85) among women exposed to non-physical IPV alone and 29.79 (95%CI: 25.18-34.41) among women with no IPV exposure. Differences between the “Both” group and each comparison group were statistically significant (p=0.02 and p<0.01, respectively). For urgent, emergent, or resuscitation-level visits (CTAS 3-1), incidence rates were directionally higher among women exposed to both forms of IPV (128.98; 95%CI: 101.71-156.24) versus non-physical IPV alone (98.24; 95%CI: 82.70-115.17) and no IPV (99.96; 95%CI: 90.83-108.48), though these differences were not statistically significant.

These findings indicate that IPV, particularly when involving physical violence, shapes long-term patterns of ambulatory care use. Differences reflect how survivors navigate healthcare systems structured by trauma, stigma, and access constraints, underscoring the need for trauma– and violence-informed care approaches.

## Introduction

Nearly 40% of Canadian women experience intimate partner violence (IPV) in their lifetime (Cotter, 2021). The World Health Organization (World Health Organization, 2013, 2021) highlights this as an ongoing and pervasive issue affecting women worldwide. IPV is not just a personal issue, it has has serious, measurable consequences for health systems (Bonomi et al., 2009; Dusing et al., 2025; Hisasue et al., 2024; William et al., 2022). This study focuses on the impact of IPV on the use of ambulatory care services among Canadian women.

This study is informed by the health equity framework (HEF; Peterson et al., 2021), which posits that health outcomes are shaped by fair access to the resources and opportunities needed for well-being. This perspective encourages researchers to look beyond individual behaviors to instead address the systemic roots of inequity. We apply this lens to understand IPV not as an isolated personal issue, but as a critical social determinant of health that creates profound health inequities by impeding access to resources and opportunities (described forthwith). This framework clarifies how structural barriers and social conditions interact to shape survivors’ long-term patterns of healthcare use, including their engagement with both physical and non-physical (e.g., coercive control, verbal and emotional abuse) forms of violence.

The HEF recognizes that systemic inequities often manifest as direct physical and psychological harm. Physical violence is associated with musculoskeletal injuries, soft tissue trauma, and repetitive traumatic brain injuries (Gosangi et al., 2021; Gujrathi et al., 2022; Valera et al., 2019). These injuries can result in chronic pain and, over time, contribute to elevated risk of neurodegenerative, cardiovascular, and mental health conditions (Iverson et al., 2017; Izzy et al., 2023). In parallel, psychological trauma can persist long after the relationship ends and is itself a key predictor of frequent ambulatory care use (Azar et al., 2020; Fedovskiy et al., 2008). Importantly, IPV is often cyclical, with past victimization increasing the risk of future exposure (Kuijpers et al., 2012), which may compound these long-term health effects. Recent global meta-analysis (Spencer et al., 2023) underscores these associations, the cumulative toll of IPV on wellbeing, and the need for more research in this area. While the link between physical violence and health is well-established, the impact of non-physical IPV is less understood. Growing evidence suggests it can have long-term health consequences comparable to those of physical violence, including an increased risk of depression, anxiety, chronic pain, and migraines (Coker, 2000; Pico-Alfonso et al., 2006), though these pathways remain understudied (Stubbs & Szoeke, 2022).

We have thus far seen how the health consequences of both physical and non-physical IPV may increase survivors’ health services needs. At the same time, the social context of IPV may present significant barriers to accessing these services. Most directly, women may be prevented from seeking care by controlling partners (Stark, 2007). Furthermore, IPV is connected to isolation and loneliness (Boyda et al., 2015; Goodman & Epstein, 2022) and to disruptions within support networks (Dias et al., 2019); factors that may delay care-seeking or complicate recovery (Wright et al., 2022). Altogether, IPV creates a challenging paradox for health systems: it generates the need for services among survivors while simultaneously creating barriers to access.

This paradox is particularly salient in Canada, where chronic under-resourcing contributes to persistent primary care shortages and forces a greater reliance on ambulatory care services (Ohle et al., 2017; Rahman et al., 2023). It is therefore crucial to understand the specific drivers of ambulatory care demand in this context, including the long-term impact of IPV. In 2022-2023, unscheduled ED visits in Canada exceeded 15 million (Canadian Institute for Health Information, 2024), the highest recorded number since such statistics were first compiled. During this period in Ontario, Canada’s most populous province, admitted patients waited an average of 23.3 hours for an inpatient bed, and fewer than one in four were admitted within the system’s 8-hour target. These delays have fatal results (Guttmann et al., 2011). Therefore, investing in upstream programs to prevent IPV is a critical strategy for strengthening health system resilience and advancing health equity.

While the extant literature suggests a link between IPV and healthcare use, its application for health system planning is hampered by small sample sizes (Pico-Alfonso et al., 2006), reliance on clinical samples (Gosangi et al., 2021; Gujrathi et al., 2022), and a focus on specific populations (Fedovskiy et al., 2008). Further, these studies are cross-sectional and are not designed to assess how IPV exposure influences long-term demand. Informed by the HEF (Peterson et al., 2021), this study aims to address these gaps, and responds to the call for more longitudinal studies in this area (Spencer et al., 2023). Drawing on a population-based cohort linked to ten years of longitudinal administrative health data in Ontario, this study examines whether past IPV exposure is associated with differences in ambulatory care use, including differences by IPV subtype.

## Methods

### Study Setting and Design

This cohort study utilized baseline data from the Neighborhood Effects on Health and Wellbeing (NEHW) study, conducted in Toronto, Canada between 2009-2011 (O׳Campo et al., 2015). NEHW used a cross-sectional design and multistage probability sampling to recruit 2,412 participants aged 25-64 who could communicate in English and had lived in their neighborhood for at least six months. Participants were sampled from 87 census tracts across 47 neighborhoods, selected to capture Toronto’s cultural and ethnic diversity. Of the original 2,412 NEHW participants, the cohort for this current study comprised N=1,094 NEHW participants who (1) provided a valid Ontario Health Insurance Plan (OHIP) number at the time of survey participation, (2) who answered the questions pertaining to IPV (detailed below), and (3) who identified as women in NEHW. The selection of NEHW participants into our analytic sample are described in Figure 1.

**Figure 1:**
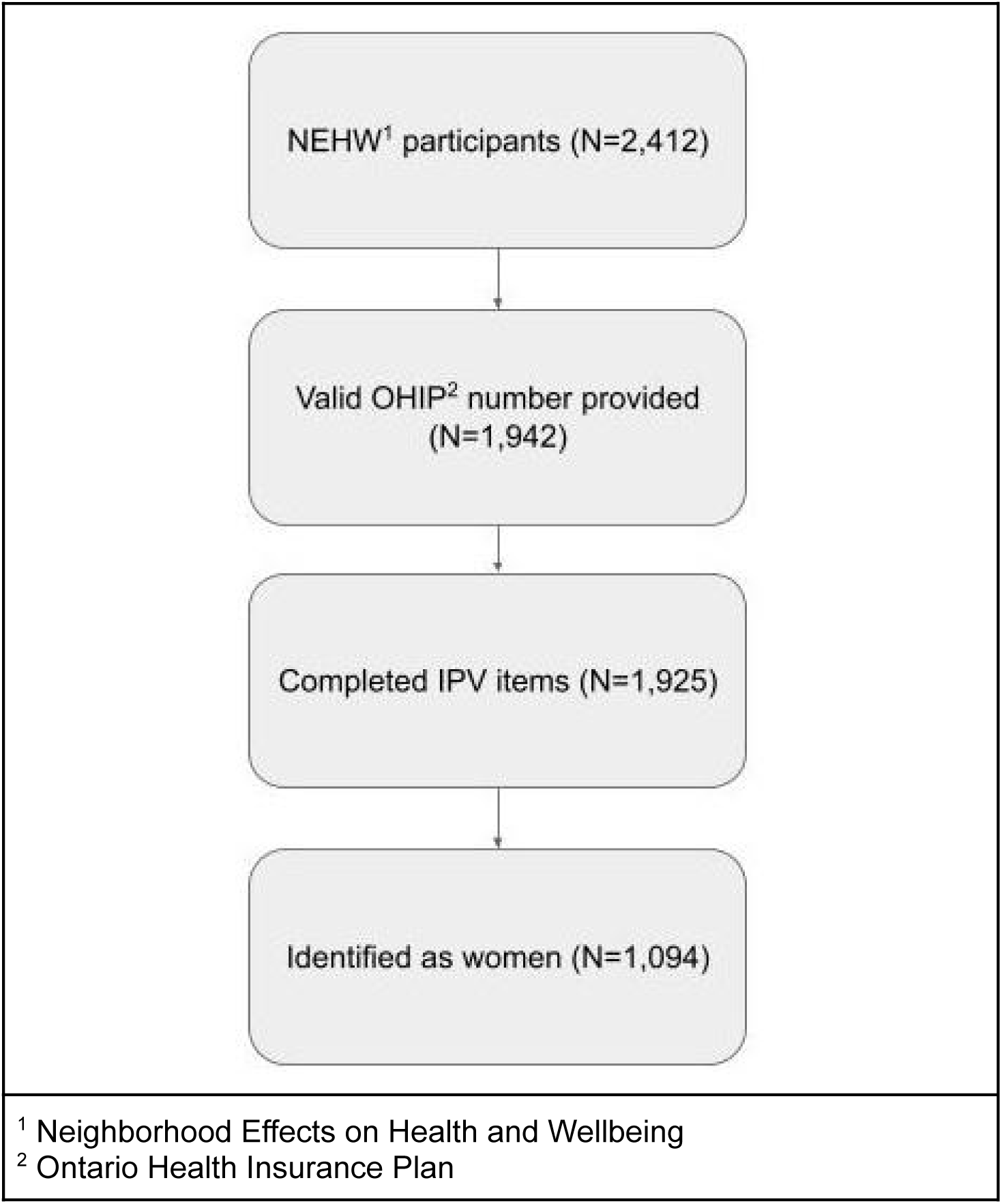
Flow of NEHW1 Participants into the Analytic Sample.

### Data Linkage

Respondents’ interactions with ambulatory care were captured through the National Ambulatory Care Reporting System (NACRS) database (Canadian Institute for Health Information, n.d.). NACRS records hospital– and community-based ambulatory care, including day surgeries, outpatient visits, and emergency department presentations. Due to NACRS’ comprehensive coverage, loss to follow-up is not a major concern in our study since it primarily occurs if individuals lose OHIP eligibility, either via moving out of province or death. Mean follow-up duration and proportion lost to follow-up are reported in our descriptive results.

IC/ES (formerly the Institute for Clinical Evaluative Sciences) is the entity authorized by the Ontario government to manage provincial healthcare data (Schull et al., 2019). IC/ES was contracted to link respondents’ OHIP records with NACRS, covering all ambulatory care interactions from their NEHW interview date to March 31, 2020. The final linked dataset for this study is housed with IC/ES to ensure compliance with Ontario’s privacy laws.

### Exposure

The main exposure of this study was exposure to IPV prior to 10 years of participation in the NEHW study, categorized by the nature of violence. In the survey, participants were asked if they had experienced any IPV in the last 10 years based on the Hurt, Insult, Threaten, Scream (HITS) scale–a short IPV screening tool (Sherin et al., 1998). If a participant answered yes to any of the HITS questions and an additional question about whether their partner imposed restrictions on their actions (Yakubovich et al., 2021), they were then asked a series of questions on their experience of physical and non-physical IPV within the past two years. Physical IPV was assessed using a 7-item version of the Physical Partner Abuse Scale (PASPH), inquiring about frequencies of being hit, shoved, beaten, or physically forced to have sex by their partner (Attala et al., 1994). Non-physical IPV was assessed using a 13-item version of the Non-Physical Partner Abuse Scale (PASNP), asking about frequencies of emotional abuse, restriction of actions, coercion, and jealousy (Attala et al., 1994). Each scale was scored by summing item responses. Both were dichotomized to reflect exposure to physical or non-physical IPV, using cut-offs of 2 for the PASPH and 15 for the PASNP (Attala et al., 1994). These recommendations were intended to balance achieving high sensitivity (98.9%) and specificity (88%), while ensuring low rates of false positives (12.0%) and negatives (2.2%) (Attala et al., 1994). In this study, IPV exposure was categorized into three groups: ‘None’ (screened negative on HITS), ‘Both Physical and Non-physical IPV’ (met cutoffs on both PASPH and PASNP), and ‘Non-physical IPV alone’ (met the PASNP cutoff but not PASPH). These categories reflect the rare occurrence (<0.4%) of meeting the PASPH cutoff but not PASNP.

### Outcome

The primary outcome was ambulatory care presentations over the study period, that is, from the time of NEHW survey participation until March 31, 2020. We analyzed this outcome in three ways: First, we examined the cumulative count of all ambulatory care presentations for each participant over the study period. Next, among those who had at least one ambulatory care presentation over the study period, we examined patterns of ambulatory care use by urgency and mode of arrival. The urgency of ambulatory care was examined by the Canadian Triage and Acuity Scale (CTAS) level categorized into two groups: CTAS levels 1 (non-urgent) and 2 (less urgent), and in the other group, CTAS levels 3 (urgent), 4 (emergent), and 5 (resuscitation). Mode of arrival was categorized as walk-in or ambulance.

### Covariates

Covariates in the fully-adjusted regression models (described below) adjusted for likely confounders at baseline (i.e., in NEHW), including age (continuous), current level of morbidity via the Charlson Comorbidity Index (CCI), current marital status (married/unmarried), childbearing history (0 children [nulliparous], 1-2, or more than 2), foreign-born status (yes/no), employment status (employed/not employed), and neighborhood material deprivation quintiles (Matheson et al., 2012).

Household income was categorized as (1) less than CA$50,000 or not stated, and (2) at least CA$50,000. On the original NEHW survey, income was collected in nine categories: one for not stated, six below CA$50,000, one between CA$50,000–73,999, and one at or above CA$74,000. Because the 2011 Ontario median household income (CA$66,358; Statistics Canada, 2019) fell within the CA$50,000–73,999 category, we selected the closest available category boundary closest to the provincial median (≤CA$50,000 vs >CA$50,000).

The CCI was included as a measure of the burden of chronic and comorbid conditions and is predictive of increased healthcare use (Charlson et al., 2022; Sundararajan et al., 2004). Neighborhood-level material deprivation was captured through quintiles from the Ontario Marginalization Index (Matheson et al., 2012). It was calculated using census data related to income, housing quality, and educational attainment and measures the extent of difficulties in meeting essential needs within a given area. Material deprivation at the neighborhood level has been associated with differential patterns of healthcare utilization (Zhang et al., 2020) and IPV risk (Bonomi et al., 2014; Cunradi et al., 2000; Kirst et al., 2015; Yakubovich et al., 2021).

### Descriptive and Statistical Analyses

In all descriptive and statistical analyses, estimates were considered statistically significant at the 0.05 level. Respondents’ characteristics at the time of survey participation were summarized using frequencies and proportions for categorical variables and means with measures of spread (standard deviation) for continuous variables. These characteristics were stratified by category of IPV exposure within 10 years of survey participation (i.e., None, both physical/sexual and non-physical IPV, and non-physical IPV alone). The distribution of the top 10 most common ICD-10 diagnosis codes for ambulatory care was examined over the full study follow-up period from the time of survey participation until March 31, 2020. Frequencies and proportions of the top-10 codes were calculated overall and stratified by baseline IPV status. Pearson’s chi-square test assessed whether the distribution of these top 10 reasons for ambulatory care differed significantly across the IPV exposure category.

The association between IPV exposure (or lack thereof) and cumulative ambulatory care use over the study period was analyzed using negative binomial regression, and incorporated follow-up time as an offset variable to account for varying durations between NEHW survey completion and the end of the study period. A zero-inflated model was not used because the proportion of zero counts was minimal (see Table 1 and Results), and the observed mean-variance relationship reflected standard overdispersion rather than zero inflation. Then, for participants with at least one ambulatory care interaction, we conducted additional analyses examining the association between IPV and both the urgency of care (based on CTAS levels) and the mode of arrival (walk-in vs. ambulance). For each outcome, regression models adjusted for the covariates detailed previously. To ensure that estimated variances are robust to model misspecification, all models were fitted with Huber-White sandwich estimators (Freedman, 2006). Crude and age-adjusted rates are presented as supplementary materials.

**Table 1:** Characteristics of Study Sample at the Time of Participation in NEHW by IPV Status.

|  | No IPV Reported within 10-years of Survey Participation |  | Non-physical IPV Alone |  | Both Physical and Non-physical IPV |  | Overall |  | p-value* |
| --- | --- | --- | --- | --- | --- | --- | --- | --- | --- |
|  | *Frequency* | *Row %* | *Frequency* | *Row %* | *Frequency* | *Row %* | *Frequency* | *Row %* |  |
| **Distribution by IPV Status** | 735 | 67.15% | 221 | 20.20% | 138 | 12.61% | 1,094 | 100.00 | N/A |
|  | *Mean* | Standard Deviation, SD | *Mean* | SD | *Mean* | SD | *Mean* | SD |  |
| **Number of Ambulatory Care Visits over Study Period** |  |  |  |  |  |  |  |  | <0.001 |
|  | 3.37 | 4.59 | 3.32 | 4.85 | 5.57 | 8.11 | 3.64 | 5.26 |  |
| **Follow-up Time (Days)** |  |  |  |  |  |  |  |  | <0.001 |
|  | 3418.98 | 479.61 | 3527.05 | 439.32 | 3535.94 | 391.74 | 3455.57 | 464.01 |  |
| **Age at Survey Participation** |  |  |  |  |  |  |  |  | <0.001 |
|  | 50.10 | 10.46 | 47.38 | 10.09 | 44.08 | 10.83 | 48.79 | 10.63 |  |
| **Charlson Comorbidity Index** |  |  |  |  |  |  |  |  | <0.001 |
|  | 2.30 | 2.25 | 1.91 | 2.07 | 1.66 | 1.83 | 2.14 | 2.17 |  |
|  | *Frequency* | *Column %* | *Frequency* | *Column %* | *Frequency* | *Column %* | *Frequency* | *Column %* |  |
| **Lost to Follow-up<sup>+</sup>** |  |  |  |  |  |  |  |  | 0.93 |
| No | 692 | 94.15% | 208 | 94.12% | 131 | 94.93% | 1031 | 94.24 |  |
| Yes | 43 | 5.85% | 13 | 5.88% | 7 | 5.07% | 63 | 5.76% |  |
| **Married** |  |  |  |  |  |  |  |  | <0.001 |
| No | 293 | 39.86% | 66 | 29.86% | 81 | 58.7% | 440 | 40.22% |  |
| Yes | 442 | 60.14% | 155 | 70.14% | 57 | 41.3% | 654 | 59.78% |  |
| **Number of Children** |  |  |  |  |  |  |  |  | 0.01 |
| None | 261 | 35.51% | 53 | 23.98% | 55 | 39.86% | 369 | 33.73% |  |
| 1-2 | 354 | 48.16% | 120 | 54.3% | 64 | 46.38% | 538 | 49.18% |  |
| 3+ | 120 | 16.33% | 48 | 21.72% | 19 | 13.77% | 187 | 17.09% |  |
| **Foreign Born** |  |  |  |  |  |  |  |  | 0.07 |
| No | 456 | 62.04% | 155 | 70.14% | 84 | 60.87% | 695 | 63.53% |  |
| Yes | 279 | 37.96% | 66 | 29.86% | 54 | 39.13% | 399 | 36.47% |  |
| **Employment** |  |  |  |  |  |  |  |  | 0.53 |
| Unemployed | 220 | 29.93% | 70 | 31.67% | 36 | 26.09% | 326 | 29.8% |  |
| Employed | 515 | 70.07% | 151 | 68.33% | 102 | 73.91% | 768 | 70.2% |  |
| **Household Income** |  |  |  |  |  |  |  |  | <0.001 |
| Less than \$50,000 Or Not Stated | 312 | 42.45% | 86 | 38.91% | 78 | 56.52% | 476 | 43.51% | |
| At Least \$50,000 | 423 | 57.55% | 135 | 61.09% | 60 | 43.48% | 618 | 56.49% | |
| **Neighborhood-level Material Deprivation Quintile^** |  |  |  |  |  |  |  |  | 0.01 |
| 1 (Least Materially Deprived | 164 | 22.31% | 38 | 17.19% | 13 | 9.42% | 215 | 19.65% |  |
| Neighborhoods) |  |  |  |  |  |  |  |  |  |
| 2 | 134 | 18.23% | 34 | 15.38% | 21 | 15.22% | 189 | 17.28% |  |
| 3 | 110 | 14.97% | 30 | 13.57% | 19 | 13.77% | 159 | 14.53% |  |
| 4 | 150 | 20.41% | 52 | 23.53% | 38 | 27.54% | 240 | 21.94% |  |
| 5 (Most<br>Materially<br>Deprived<br>Neighborhoods) | 177 | 24.08% | 67 | 30.32% | 47 | 34.06% | 291 | 26.6% |  |
| Note:<br>* p-values derived from chi-square tests for categorical variables and ANOVA for means to assess differences in the distribution of characteristics across the three IPV exposure groups.<br>+ Respondents who died or lost OHIP eligibility before the end of the study period.<br>^ Neighborhood deprivation based on quintiles provided by the 2011 Ontario Marginalization Index. |  |  |  |  |  |  |  |  |  |

Using the estimated models, we calculated incidence rates per 100,000 person-years for each outcome and IPV status categories. For the cumulative count outcome, these rates represent the number of ambulatory care visits per 100,000 person-years. For the urgency of care and mode of arrival outcomes, the rates indicate the number of visits per 100,000 person-years for each specific category (e.g., less urgent visits, ambulance arrivals) among those with at least one ambulatory care interaction.

After estimating incidence rates per 100,000 person-years for each IPV status category, we conducted pairwise comparisons with the Bonferroni correction for multiple comparisons. This analysis compared the estimated incidence rates between each pair of IPV status categories (i.e., None vs. Both physical/sexual and non-physical IPV, None vs. non-physical IPV alone, and non-physical IPV alone vs. Both physical/sexual and non-physical IPV), calculating the differences in rates and assessing their statistical significance. The Bonferroni correction was applied to adjust p-values, reducing the risk of Type I errors associated with multiple comparisons and to keep the overall threshold for statistical significance at alpha = 0.05.

### Sensitivity Analyses

We conducted several sensitivity analyses to evaluate the robustness of our findings. Since chronic conditions influence healthcare utilization, a supplementary analysis was conducted on the subset with CCI values of 2 or less (representing the median CCI and the healthiest individuals in our sample). To evaluate the impact of exposure recency, models were refitted using only those with IPV exposure within 2 years of participating in NEHW. To do so, we refitted the models using a modified exposure classification: the ‘None’ group remained unchanged (No IPV reported within 10 years of NEHW participation), but we reclassified the other two groups (’Both physical/sexual and non-physical IPV’ and ‘Non-physical IPV alone’) to include only those who experienced IPV within 2 years of NEHW participation. All sub-analyses results are available in the supplementary materials.

## Results

### Distribution of Characteristics

Table 1 reports the distribution of covariates by IPV category (None, Non-physical IPV alone, and Both Physical and Non-physical IPV). Differences in covariate distributions were assessed using one-way ANOVA for comparing means of continuous variables and chi-square tests for categorical variables. Of the 1,094 women in our study, 32.81% reported exposure to some form of IPV within 10 years prior to NEHW participation: 12.61% experienced both physical and non-physical IPV, while 20.20% experienced non-physical IPV alone.

The overall mean follow-up duration was 3455.57 days (SD=464.01). Mean follow-up time differed by IPV category (p<0.01), with those in the “None” group followed-up the shortest follow-up time (mean=3418.98, SD=479.61) and those in the “Both” group the longest (mean=3455.57, SD=464.01). Although mean follow-up time varied between IPV categories, the overall proportion of those lost to follow-up (i.e., those who died or otherwise lost OHIP coverage) before the end of the study period was 5.76%, with little variation by category (p=0.93). Having examined the distribution of characteristics across IPV categories, we next analyzed the most common reasons for ambulatory care visits.

Women in the “Both Physical and Non-physical IPV” (henceforth, the “Both” group) group appeared to be younger and healthier than women who reported either no IPV (the “None” group) or only non-physical IPV (the “Non-physical Alone” group) within ten years of participating in NEHW. Specifically, women in the “Both” group were younger (mean=44.08, standard deviation [SD]=10.83), compared to those in the “None” group (mean=50.10, SD=10.46) and “Nonphysical Alone” group (mean=47.38, SD=10.09) (p-value, p<0.01). Women in the “Both” group had the lowest mean CCI (mean=1.66, SD=1.83), compared to the “None” (mean=2.30, SD=2.20) and “Nonphysical Alone” (mean=1.91, SD=2.07) groups (p<0.01).

Women in the “Both” group lived in houesholds with the lowest incomes, with 56.52% living in households earning less than CA$50,000 (or not stated), compared to 42.45% in the “None” and 38.91% in the “Nonphysical Alone” groups (p<0.01). Additionally, women in the “Both” group were more likely to live in the most materially deprived neighborhoods, with 61.6% residing in the lowest two quintiles of neighborhood material deprivation, compared to 44.49% of women in the “None” group.

Significant associations were found between IPV exposure and both marital status at time of completion of the NEHW and number of children (p<0.01 for both). Marital status at the time of participation in NEHW varied notably across IPV categories. Women in the “Both” group were least likely to be currently married (41.3%), compared to the “None” (60.14%) and “Nonphysical Alone” (70.14%) groups.

The distribution of the number of children also differed significantly among IPV categories (p=0.01). Women in the “Nonphysical Alone” group were most likely to have children, with only 23.98% nulliparous women, compared to 35.51% of women in the “None” and 39.86% of the “Both” groups. The proportion of women with 1-2 children was highest in the “Nonphysical Alone” group (54.3%), compared to 48.16% in the “None” group and 46.38% in the “Both” group. Interestingly, the “Nonphysical Alone” group also had the highest proportion of women with 3 or more children (21.72%), compared to 16.33% in the “None” group and 13.77% in the “Both” group.

### Main Presenting Complaints

Table 2 presents the ICD-10-CA codes for the top 10 most common main complaints at ambulatory care presentation. The most common complaints across all groups were R10.4 “Other and unspecified abdominal pain” (23.37% of visits overall), R07.4 “Unspecified chest pain” (21.4%), and N39.0 “urinary tract infection, site not specified” (11.56%). Notably, abdominal pain was more prevalent in the IPV-exposed groups, particularly in the “Both” group (31.22%) compared to the “None” group (19.53%). Chest pain was also slightly more common in the “Both” group (24.87%) compared to others. Other frequent complaints included open wounds, other chest pain, gastroenteritis, vertigo, syncope, headache, and cellulitis. Urinary tract infections were slightly less common in the “Both” group compared to others (“Both” – 9.52% vs. “None” – 12.31% and “Non-physical Alone” – 11.76%). Despite these apparent differences between IPV categories, they did not reach statistical significance, having a chi-squared p-value of 0.15.

**Table 2:** Distribution of Top 10 ICD-10 Codes for Main Complaint in Ambulatory Care Over the Follow-Up Period by IPV Status at Time of Survey Participation.

|  |  | No IPV |  | Non-physical IPV Alone |  | Both Physical and Non-physical IPV |  | Overall |  |
| --- | --- | --- | --- | --- | --- | --- | --- | --- | --- |
| Code Title | ICD-10-CA Code | N | % | N | % | N | % | N | % |
| Other and unspecified abdominal pain | R10.4 | 92 | 19.53% | 39 | 25.49% | 59 | 31.22% | 190 | 23.37% |
| Chest pain, unspecified | R07.4 | 97 | 20.59% | 30 | 19.61% | 47 | 24.87% | 174 | 21.40% |
| Urinary tract infection, site not specified | N39.0 | 58 | 12.31% | 18 | 11.76% | 18 | 9.52% | 94 | 11.56% |
| Open wound of thumb without damage to nail | S61.00 | 37 | 7.86% | 11 | 7.19% | 7 | 3.70% | 55 | 6.77% |
| Other chest pain | R07.3 | 37 | 7.86% | 13 | 8.50% | 12 | 6.35% | 62 | 7.63% |
| Gastroenteritis and colitis of unspecified origin | A09.9 | 34 | 7.22% | 1-5* | 0.65-3.27% | 10-14* | 5.29-7.41% | 50 | 6.15% |
| Vertigo | R42 | 30 | 6.37% | 9 | 5.88% | 10 | 5.29% | 49 | 6.03% |
| Syncope and collapse | R55 | 32 | 6.79% | 8 | 5.23% | 10 | 5.29% | 50 | 6.15% |
| Headache | R51 | 30 | 6.37% | 11 | 7.19% | 9 | 4.76% | 50 | 6.15% |
| Cellulitis of lower limb | L03.11 | 24 | 5.10% | 8-12* | 5.23-7.84% | 1-5* | 0.53-2.65% | 39 | 4.80% |
| Total |  | 471 | 100.00% | 153 | 100.00% | 189 | 100.00% | 813 | 100.00% |

Note:
Pearson Chi-squared Value(18) = 32.46; p-value = 0.15
\* Exact frequency suppressed due to cell counts falling below IC/ES vetting requirements.

### Association of IPV with Ambulatory Care Use

The overall mean cumulative ambulatory care interactions was 3.64 (SD=5.26) (Table 1), with significant differences across IPV categories (p<0.01). The “Both” group had the highest mean at 5.57 (SD=8.11). Using negative binomial regression adjusted for follow-up time and covariates listed in our methods section, we estimated incidence rates per 100,000 for ambulatory care by IPV status (Table 3a). The incidence rates were 145.16 (95% CI: 111.69-178.64) for the “Both” group, 90.37 (95% CI: 73.58-107.16) for the “Nonphysical Alone” group, and 98.74 (95% CI: 88.93-108.56) for the “None” group. Pairwise comparisons (Table 3b) revealed significant differences between the “Both” group and both the “None” group (contrast=0.39; p=0.01) and the “Nonphysical Alone” group (contrast=0.47; p=0.01). Although the “Nonphysical Alone” group appeared to have lower incidence rates per 100,000 (90.37; 95%CI: 73.58-107.16) than the “None” group (98.74; 95%CI: 88.93-108.56), these differences did not reach statistical significance (contrast=-0.09; p=1.00).

**Table 3a:**
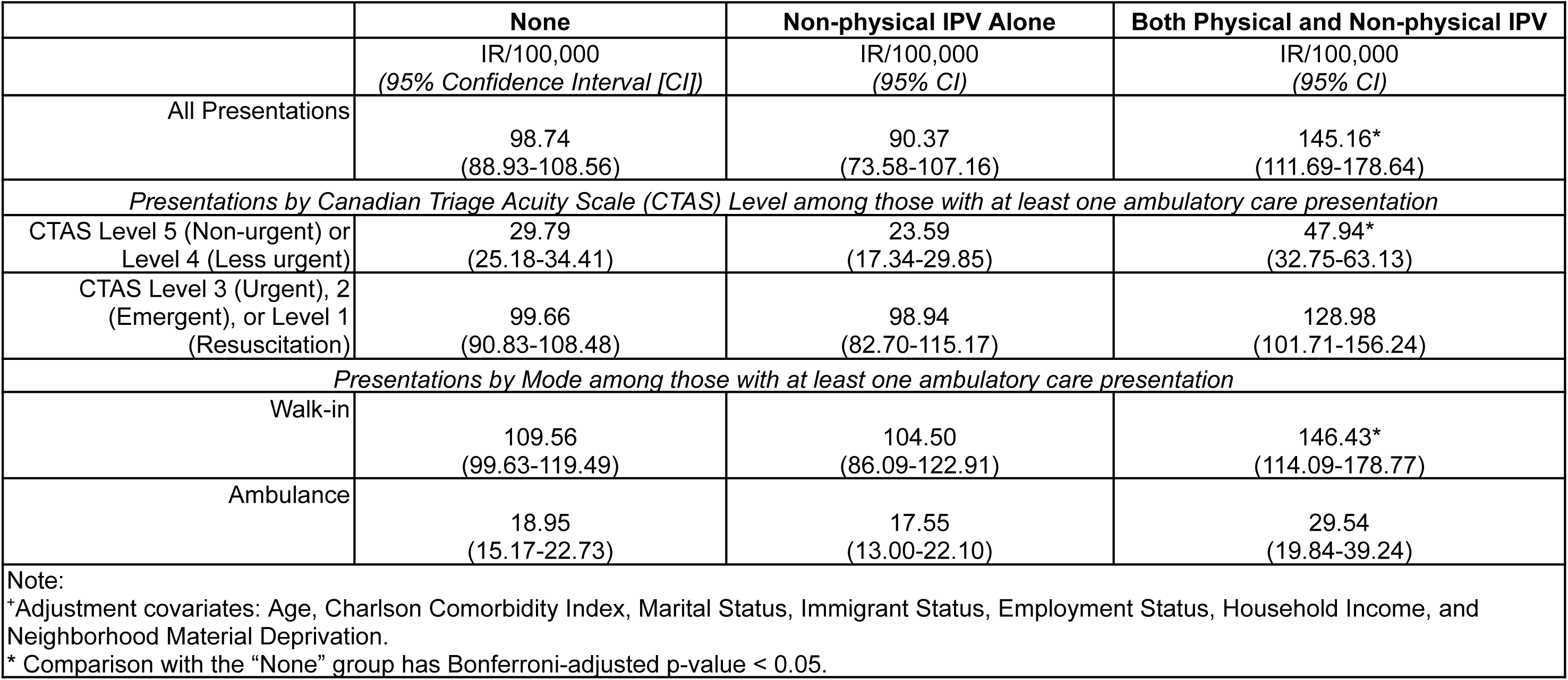
Fully-adjusted + Incidence Rates per 100,000 (IR/100,000) of Ambulatory Care Presentations Over the Study Period (2009-2020) by IPV Status Over the Study Period (2009-2020).

**Table 3b:**
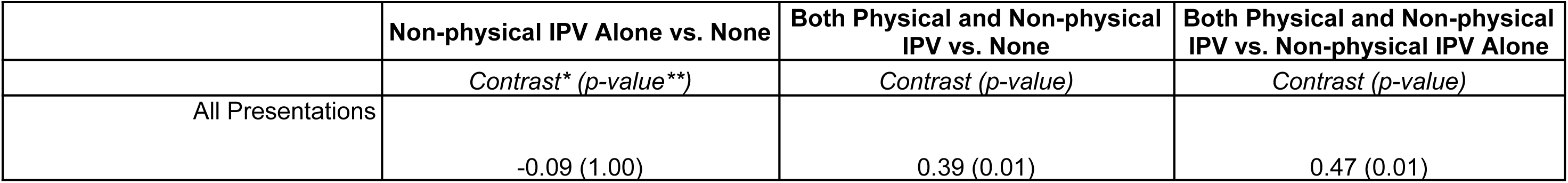
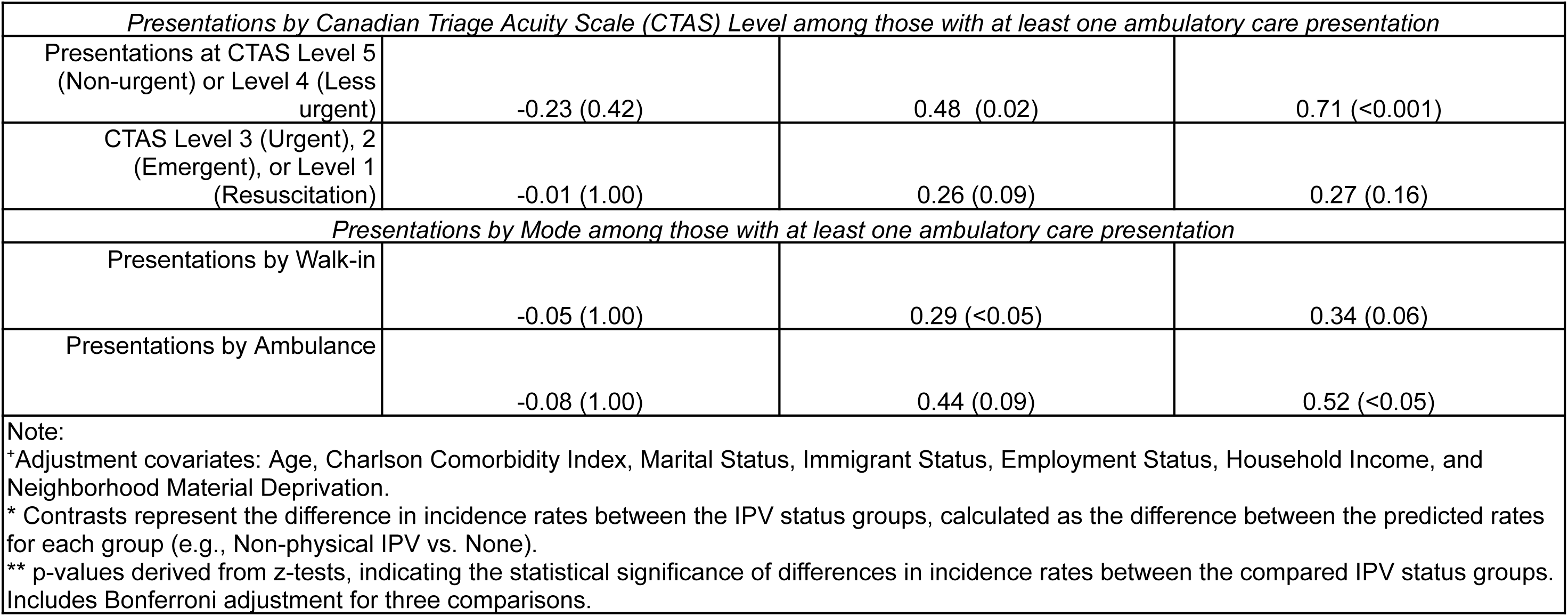
Comparison of fully-adjusted+ incidence rates per 100,000 (IR/100,000) of Ambulatory Care Presentations Over the Study Period (2009-2020) by IPV Status.

We then compared incidence rates of ambulatory care presentations by triage acuity. Residents exposed to both physical and non-physical IPV had higher rates of urgent and emergent visits (CTAS 3–1) compared with those reporting non-physical IPV alone or no IPV. Although these differences did not reach statistical significance, the pattern suggests that individuals experiencing both forms of IPV are more likely to present with higher-acuity concerns. In contrast, for non-urgent visits (CTAS 4–5), rates were significantly higher in the “Both” group than in either comparison group (p=0.02 vs. “None”; p<0.01 vs. “Non-physical IPV Alone”).

Finally, among those who had at least one ambulatory care presentation, we examined presentations by mode (i.e., presentations by either walk-in or ambulance). First, the incidence rates per 100,000 for presentations by walk-in were 146.43 (95%CI: 114.09-178.77), 104.50 (95%CI: 86.09-122.91), 109.56 (95%CI: 99.63-119.49) for the “Both,” “Nonphysical IPV Alone,” and “None” groups respectively. Only the difference between the “Both” and “None” groups were significant (contrast=0.29; p<0.05). The incidence rates per 100,000 for presentations by ambulance were 29.54 (95%CI: 19.84-39.24), 17.55 (95%CI: 13.00-22.10), 18.95 (95%CI: 15.17-22.73) for the “Both,” “Nonphysical IPV Alone,” and “None” groups respectively, and again, only the difference between “Both” and “None” were significant (contrast=0.52; p<0.05).

### Results of Sensitivity Analyses

In the analysis restricted to individuals with a CCI of at most one (Supplementary Tables 5 & 6), exposure to both physical and non-physical IPV was associated with a higher rate of ambulatory care utilization (IRR: 1.70, 95%CI:1.15-2.50). For those living in neighborhoods with the greatest deprivation (5th Quintile) (Supplementary Table 7), exposure to both physical and non-physical IPV was associated with more frequent interactions with ambulatory care (IRR:2.07, 95%CI:1.32-3.24). Model results for the subsample whose IPV exposure occurred within 2-years of NEHW did not exhibit any significant results, likely due to the loss of statistical power. The results from these sensitivity analyses were broadly consistent with the main findings, albeit with slightly attenuated effect estimates.

## Discussion and Conclusions

### Summary of Key Findings

This study examined the long-term impact of IPV exposure on ambulatory care use among women in Toronto, Canada, using data from the NEHW study (2009–2011) linked to NACRS records through 2020. In adjusted analyses, we found that women exposed to both physical and non-physical IPV had significantly higher rates of ambulatory care use over a ∼10-year follow-up, compared to those reporting no IPV or non-physical IPV only. Among those with at least one visit, women exposed to both physical and non-physical IPV also had higher rates of less-or non-urgent care and walk-in visits. In contrast, exposure to non-physical IPV alone was not associated with increased ambulatory care use and appeared to be linked to lower utilization across most presentation types—though these differences were not statistically significant. It is important to note that differences in care use across IPV groups reflect variations in how survivors interact with health systems, not normative differences in care-seeking behavior or system contact, which we will elaborate in conclusion.

### Comparison with Previous Research

Our findings that women exposed to both physical and non-physical IPV have higher rates of ambulatory care use compared to those reporting no violence align with existing literature. Bonomi et al. (2009) reported similar findings in their study. They observed that women exposed to IPV more than 5 years prior had 52% higher incidence of emergency department visits over the following 10-year period compared to women reporting no IPV. This effect was even more pronounced for recent exposure, with women experiencing IPV within the past 5 years showing a 68% higher risk. The same study also found increased risk of outpatient hospital visits: women with IPV exposure over 5 years ago had 15% higher risk ratios, while those with exposure within the past 5 years had 34% higher risk compared to women with no reported IPV. Similarly, Dichter et al. (2018) found that women disclosing IPV in the Veterans Health Administration had significantly higher odds of using emergency, inpatient, and mental health services, compared to those not reporting IPV. Recent evidence from Finland further reinforces this pattern, showing that IPV survivors identified through linked policy and health registers incurred mean attributable healthcare costs of 3,280 Euros over a five-year period (Hisasue et al., 2024). In contrast, studies from low– and middle-income countries (LMICs) report a strikingly different trend: women exposed to IPV are often less likely to use some health services (Leight & Wilson, 2021). This is not to say that women in LMICs need less services, and instead this pattern may be due to: (1) coercive control and restrictions on mobility, (2) that women in violent relationships in these settings are more likely to seek help from informal rather than formal channels (Kanougiya et al., 2022; World Health Organization, 2005), and (3) the lack of and the cost to use health services (World Health Organization, 2005). While these studies did not distinguish IPV by type, their findings support the general association between IPV and greater health service utilization, particularly when physical violence is involved.

These findings are consistent with the World Health Organization Pathways’ model (World Health Organization, 2013b), which posits that exposure to violence can lead to increased health service use through multiple physical, psychological, and behavioral pathways. However, these results should be interpreted with caution, as our data capture only a subset of health services—primarily ambulatory and hospital-based care—and do not include most mental health or community-based supports, where women exposed to non-physical IPV may be more likely to seek help.

In contrast, exposure to non-physical IPV alone was not associated with increased ambulatory care use and appeared to be linked to lower utilization across most presentation types—although these differences were not statistically significant. However, this finding should be interpreted with caution, as non-physical IPV may lead survivors to seek care primarily through mental health or social support services not captured within NACRS, rather than indicating reduced overall healthcare engagement.

To our knowledge, ours is the first study examining long-term ambulatory care use after non-physical IPV alone in a population-based, Canadian cohort. While we found a lower (though non-significant) use of ambulatory care over time for those exposed only to non-physical IPV, this does not imply non-physical IPV is any less serious. In fact, prior studies have found associations between this form of violence and negative health outcomes (Metheny et al., 2025; Saint-Eloi Cadely et al., 2020). Instead, it may indicate that survivors engage more with specialized mental health services that fall outside the scope of ambulatory care captured by NACRS. A 2020 study found that, among U.S. women screened positive for past-year IPV in Veterans Health Administration clinics, the adjusted rate ratio (aRR) for psychosocial (mental health) visits was 2.40 (95 % CI: 2.00-2.90), substantially higher than the rates for primary care (aRR = 1.20, 95 % CI: 1.10-1.30) and emergency department use (aRR = 1.50, 95 % CI: 1.20–1.80) (Makaroun et al., 2020).

### Strengths and Limitations

Our study has several limitations. First, IPV was only captured at the time of the survey. We couldn’t account for new IPV experiences that may have occurred later, which could lead to underestimating IPV’s long-term effects. While we attempted to track IPV exposures subsequent to NEHW participation using specific ICD-10 codes (T74.0-T74.9, X85-Y09, Z63.0) related to IPV (Olive, 2018), none of the participants in our sample received these diagnostic codes during the study period. This absence of coding may reflect the underuse in clinical practice, potentially due to the stigma associated with terms such as “abuse” or “domestic violence” (Olive, 2018), or the perceived lack of utility by clinicians given the inability to bill for non-diagnostic codes. Therefore, the “no IPV” group should be interpreted as those without reported IPV at baseline rather than as a true control group. Second, while NACRS captures all ambulatory care in Ontario, it represents only one aspect of healthcare use. Notably, it does not include allied or mental healthcare use, which prior studies have found to be related to exposure to non-physical IPV (Bonomi et al., 2009). Next, this analysis is based on a cohort sample rather than a population-based dataset, the incidence rates reflect patterns within our study population and should not be interpreted as population-level estimates. Lastly, if participants lose OHIP eligibility (either through death or emigration out of the country) or seek ambulatory care outside Canada, these interactions would not be captured in the study. However, this is unlikely to be a major source of bias. The overall proportion of participants lost to follow-up was low (<6%), and critically, this proportion did not differ significantly by IPV exposure.

Despite these limitations, we note that this study has several strengths. First, our study benefits from a long follow-up time, with an overall mean of 9.5 years (3,455 days) of observation. This extended period allows us to observe the long-term impacts of IPV on ambulatory care use. Second, due to the linkage with NACRS which automatically captures any interaction with ambulatory care, we experienced very low rates of loss to follow-up, with only 5.76% of participants lost over the study period. This high retention rate enhances the reliability and validity of our findings by minimizing potential bias from differential attrition. Third, our approach to determining IPV exposure focused on specific behaviors rather than labeling them as violent. This method may have encouraged more accurate disclosure from participants who might not have recognized or labeled their experiences as IPV. Fourth, our use of NACRS ensured comprehensive capture of ambulatory care visits anywhere in Canada, providing a complete picture of ambulatory care use regardless of where participants sought care. Finally, the consistency of results between our main analyses and sub-analyses, as well as their alignment with findings from studies in other settings, strengthens the robustness of our conclusions. This consistency suggests that our findings are not artifacts of particular analytical choices and adds to the growing body of evidence on the relationship between IPV and ambulatory care use across diverse contexts.

## Conclusion

While our findings point to clear differences in ambulatory care use between women with and without IPV exposure, we caution against interpreting these differences through the lens of surveillance or normative assumptions about care-seeking. Greater or lesser use of ambulatory care should not be read as inherently positive or negative; instead, it reflects how survivors navigate health systems shaped by trauma, stigma, and structural barriers. These variations point to different journeys through care and not to deficits in individual behavior or system contact.

Screening for IPV in healthcare settings can play a vital role, particularly for survivors who may not disclose violence elsewhere. Detection can be an important first step toward accessing supports, especially when trauma is not immediately visible as in the case of nonphysical IPV. However, the act of screening must not be mistaken for care itself. Screening practices must be trauma-informed, non-punitive, and embedded within a larger system of care that includes timely referrals, community-based supports, and mental health services. Without this infrastructure, screening may inadvertently contribute to feelings of exposure without safety, reinforcing the very dynamics of control that IPV survivors often face.

The HEF (Peterson et al., 2021) emphasizes that health inequities cannot be resolved by focusing on individual choices and behaviors alone. Our findings support this view, underlining the resource demands associated with IPV. These resource demands cannot be addressed solely through individual-level interventions. The under-resourcing of both health and social care sectors in Ontario undermines the effectiveness of IPV response efforts across the continuum. No amount of screening or triage will compensate for the lack of services that survivors need in the aftermath of violence.

To move forward, policies must fund and sustain person-centered care models that go beyond episodic medical responses. This includes not only improving IPV detection and referral systems, but ensuring that there are culturally safe, accessible, and trauma-informed services for survivors to be referred into, including mental healthcare, social supports, or long-term housing.

## Declarations

Ethics approval was granted by the Research Ethics Board of St. Michael’s Hospital (REB# 20-120). This work was supported by the Social Sciences and Humanities Research Council of Canada via an Insight grant to Nicholas Metheny (#435-2020-1410).

The data used in this study are held securely in coded form at ICES. Provincial privacy legislation and data sharing agreements prohibit ICES from making the dataset publicly available. Access may be granted to approved researchers through ICES’ virtual environment following institutional credentialing and research ethics board approval.

## Acknowledgments

This work is supported by the Li Ka Shing Knowledge Institute of Unity Health Toronto, and by ICES, which provided the linkage with NEHW to the health administrative data used in this analysis. Each aspect of the research (including conceptualization, analysis, interpretation, or writing of this study) were independently carried out by the authors.

## Supplementary Materials

**Supplementary Table 1a:** Incidence Rates per 100,000 (IR/100,000) of Ambulatory Care Presentations Over the Study Period (2009-2020) by IPV Status Over the Study Period (2009-2020).

|  |  |  |  |
| --- | --- | --- | --- |
|  | None | Non-physical IPV Alone | Both Physical and Non-physical IPV |
|  | IR/100,000<br>(95% Confidence Interval [CI]) | IR/100,000<br>(95% CI) | IR/100,000<br>(95% CI) |
| **Crude Rates** |  |  |  |
| All Presentations | 101.42<br>(91.17-111.67) | 94.19<br>(76.43-111.96) | 163.83*<br>(122.44-205.23) |
| *Presentations by Canadian Triage Acuity Scale (CTAS) Level among those with at least one ambulatory care presentation* |  |  |  |
| CTAS Level 5 (Non-urgent) or<br>Level 4 (Less urgent) | 31.44<br>(26.52-36.36) | 23.89<br>(18.06-29.72) | 52.71*<br>(35.46-69.97) |
| CTAS Level 3 (Urgent), 2<br>(Emergent), or Level 1 | 100.74<br>(91.68-109.80) | 103.42<br>(85.68-121.16) | 140.82*<br>(107.50-174.14) |
| (Resuscitation) |  |  |  |
| *Presentations by Mode among those with at least one ambulatory care presentation* |  |  |  |
| Presentations by Walk-in | 109.93<br>(99.90-119.96) | 108.39<br>(89.32-127.45) | 157.79*<br>(120.29-195.29) |
| Presentations by Ambulance | 22.52<br>(18.03-27.02) | 18.71<br>(13.73-23.69) | 35.91<br>(22.65-49.17) |
| **Age-adjusted Rates** |  |  |  |
| All Presentations | 100.55<br>(90.21-110.90) | 94.43<br>(76.65-112.20) | 169.44*<br>(126.14-212.74) |
| *Presentations by Canadian Triage Acuity Scale (CTAS) Level among those with at least one ambulatory care presentation* |  |  |  |
| CTAS Level 5 (Non-urgent) or<br>Level 4 (Less urgent) | 30.77<br>(25.83-35.72) | 23.99<br>(18.15-29.84) | 55.88*<br>(37.22-74.55) |
| CTAS Level 3 (Urgent), 2<br>(Emergent), or Level 1<br>(Resuscitation) | 100.19<br>(91.03-109.35) | 103.58<br>(85.86-121.30) | 144.03*<br>(110.27-177.78) |
| *Presentations by Mode among those with at least one ambulatory care presentation* |  |  |  |
| Walk-in | 109.60<br>(99.45-119.76) | 108.50<br>(89.43-127.56) | 159.83*<br>(121.93-197.73) |
| Ambulance | 21.28<br>(16.60-25.97) | 18.68<br>(13.81-23.55) | 40.16*<br>(24.28-56.04) |
| Note:<br>- * Comparison with the "None" group has Bonferroni-adjusted p-value < 0.05.<br>- Crude rates represent crude rates per 100,000 without any adjustment<br>- Age-adjusted represent crude rates per 100,000 adjusting only for age at time of participation in NEHW. |  |  |  |

**Supplementary Table 1b:** Comparison of incidence rates per 100,000 (IR/100,000) of Ambulatory Care Presentations Over the Study Period (2009-2020) by IPV Status.

|  | **Non-physical IPV Alone vs.**<br>**None** | **Both Physical and**<br>**Non-physical IPV vs. None** | **Both Physical and**<br>**Non-physical IPV vs.**<br>**Non-physical IPV Alone** |
| --- | --- | --- | --- |
|  | *Contrast\* (p-value\*\*)* | *Contrast (p-value)* | *Contrast (p-value)* |
| **Crude Rates** |  |  |  |
| All Presentations | -0.07 (1.00) | 0.48 (0.00) | 0.55 (<0.001) |
| *Presentations by Canadian Triage Acuity Scale (CTAS) Level among those with at least one ambulatory care presentation* |  |  |  |
| CTAS Level 5 (Non-urgent) or<br>Level 4 (Less urgent) | -0.27 (0.19) | 0.52 (0.02) | 0.79 (<0.001) |
| CTAS Level 3 (Urgent), 2<br>(Emergent), or Level 1<br>(Resuscitation) | 0.03 (1.00) | 0.33 (0.03) | 0.31 (0.12) |

| *Presentations by Mode among those with at least one ambulatory care presentation* |  |  |  |
| --- | --- | --- | --- |
| Walk-in | -0.01 (1.00) | 0.36 (0.02) | 0.38 (0.04) |
| Ambulance | -0.19 (0.82) | 0.47 (0.09) | 0.65 (0.02) |
| **Age-adjusted Rates** |  |  |  |
| All Presentations | -0.06 (1.00) | 0.52 (<0.001) | 0.58 (<0.001) |
| *Presentations by Canadian Triage Acuity Scale (CTAS) Level among those with at least one ambulatory care presentation* |  |  |  |
| Presentations at CTAS Level 5<br>(Non-urgent) or Level 4 (Less<br>urgent) | -0.25 (0.29) | 0.60 (0.01) | 0.85 (<0.001) |
| CTAS Level 3 (Urgent), 2<br>(Emergent), or Level 1 | 0.03 (1.00) | 0.36 (0.01) | 0.33 (0.08) |
| (Resuscitation) |  |  |  |
| *Presentations by Mode among those with at least one ambulatory care presentation* |  |  |  |
| Presentations by Walk-in | -0.01 (1.00) | 0.38 (0.01) | 0.39 (0.03) |
| Presentations by Ambulance | -0.13 (1.00) | 0.63 (0.02) | 0.77 (<0.001) |
| Notes:<br>- Crude rates represent crude rates per 100,000 without any adjustment<br>- Age-adjusted represent crude rates per 100,000 adjusting only for age at time of participation in NEHW.<br>- * Contrasts represent the difference in incidence rates between the IPV status groups, calculated as the difference between the predicted rates for each group (e.g., Non-physical IPV vs. None).<br>- ** p-values derived from z-tests, indicating the statistical significance of differences in incidence rates between the compared IPV status groups. Includes Bonferroni adjustment for three comparisons. |  |  |  |

**Supplementary Table 1:** Unadjusted Negative Binomial Regression Models Results for Cumulative Ambulatory Care Presentations Over the Study Period (2009-2020) by IPV Status Over the Study Period (2009-2020)

|  | None | Non-physical IPV Alone | Both Physical and Non-physical IPV |
| --- | --- | --- | --- |
| **IRR**<br>**(95% Confidence Interval [CI])** | 1.00 (Reference) | 0.93<br>(0.75-1.15) | 1.62**<br>(1.23-2.12) |
| Note:<br>* p < 0.05; ** p < 0.01; *** p < 0.0001 |  |  |  |

**Supplementary Table 3:**
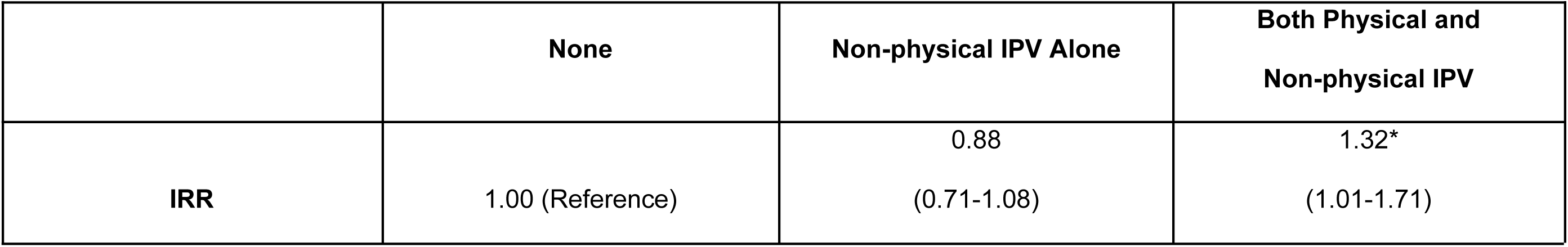
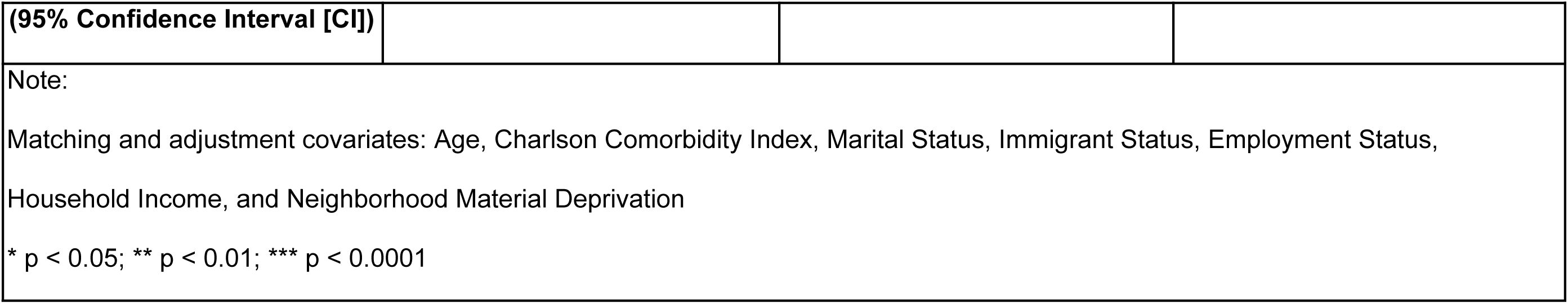
Adjusted Negative Binomial Regression Models for Cumulative Interactions with Ambulatory Care Over Study Period with Inverse Probability Weighted Cohort.

**Supplementary Table 5:**
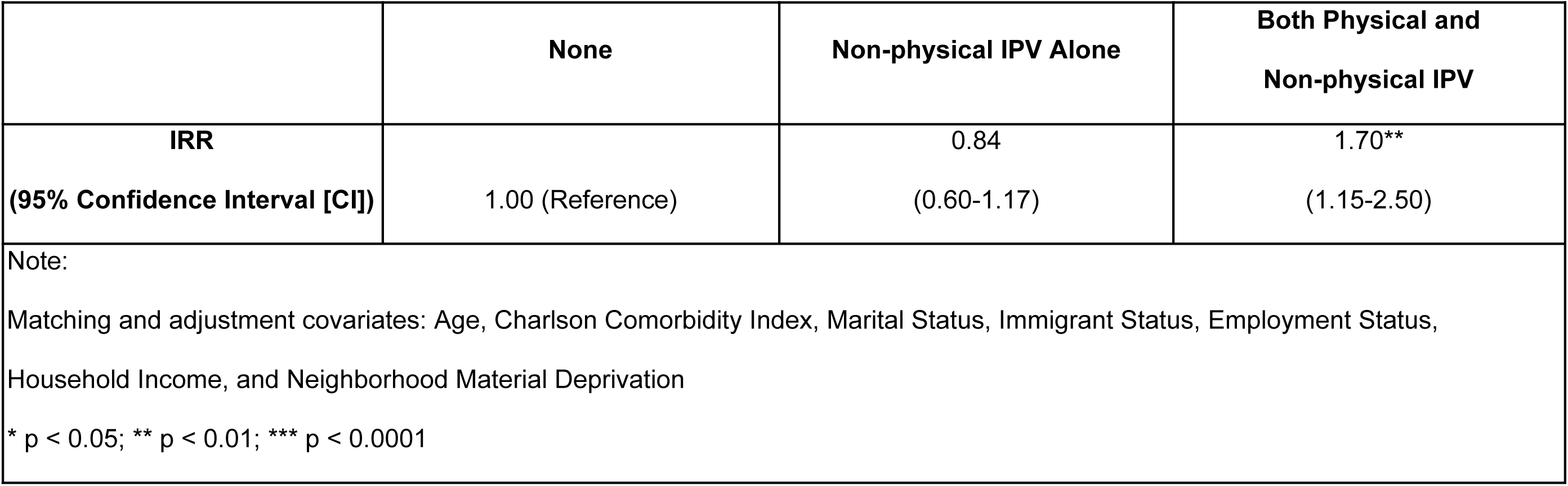
Adjusted Negative Binomial Regression Models for Cumulative Interactions with Ambulatory Care Over Study Period (Subsample with CCI <= 1)

**Supplementary Table 7:** Adjusted Negative Binomial Regression Models for Cumulative Interactions with Ambulatory Care Over Study Period (Subsample in most deprived neighborhoods [Deprivation Quintile = 5])

|  | **None** | **Non-physical IPV Alone** | **Both Physical and**<br>**Non-physical IPV** |
| --- | --- | --- | --- |
| **IRR**<br>**(95% Confidence Interval [CI])** | 1.00 (Reference) | 0.92<br>(0.64-1.33) | 2.07**<br>(1.32-3.24) |

Note:
Matching and adjustment covariates: Age, Charlson Comorbidity Index, Marital Status, Immigrant Status, Employment Status, Household Income, and Neighborhood Material Deprivation
\* $p < 0.05$ ; \*\* $p < 0.01$ ; \*\*\* $p < 0.0001$

**Supplementary Table 9:** Adjusted Negative Binomial Regression Models for Cumulative Interactions with Ambulatory Care Over Study Period (Subsample in least deprived neighborhoods [Deprivation Quintile = 1])

|  | None | Non-physical IPV Alone | Both Physical and<br>Non-physical IPV |
| --- | --- | --- | --- |
| **IRR**<br>**(95% Confidence Interval [CI])** | 1.00 (Reference) | 0.84<br>(0.57-1.25) | 0.63<br>(0.33-1.22) |
| Note:<br>Matching and adjustment covariates: Age, Charlson Comorbidity Index, Marital Status, Immigrant Status, Employment Status,<br>Household Income, and Neighborhood Material Deprivation<br>* $p < 0.05$; ** $p < 0.01$; *** $p < 0.0001$ | | | |

**Supplementary Table 11:**
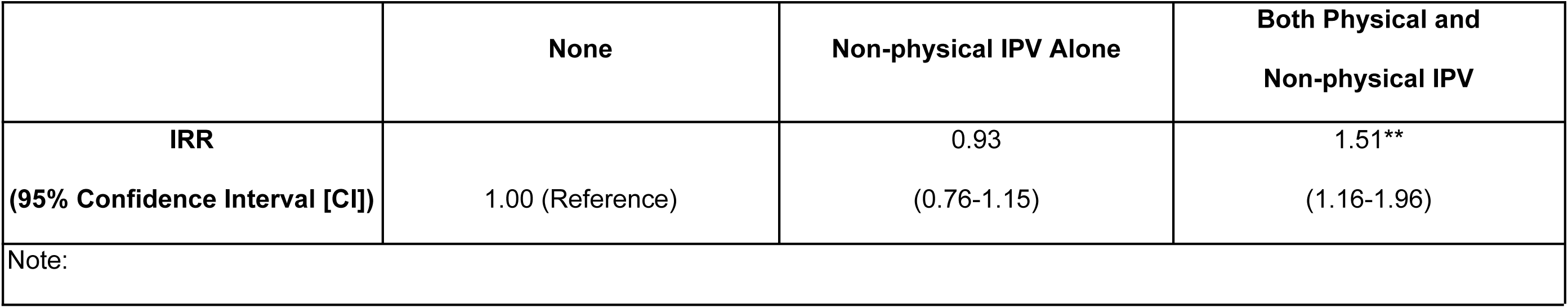
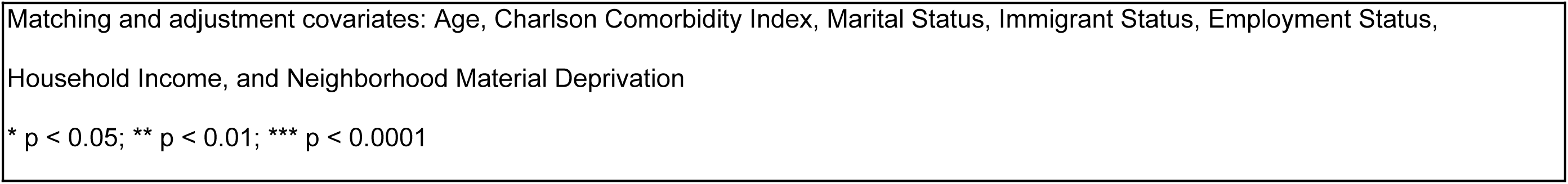
Adjusted Negative Binomial Regression Models for Cumulative Interactions with Ambulatory Care Over Study Period (Model without Neighborhood Material Deprivation)

|  |  |  |  |
| --- | --- | --- | --- |
|  | None | Non-physical IPV Alone | Both Physical and<br>Non-physical IPV |
| **IRR**<br>**(95% Confidence Interval [CI])** | 1.00 (Reference) | 0.93<br>(0.76-1.15) | 1.51**<br>(1.16-1.96) |
| Note: |  |  |  |

Matching and adjustment covariates: Age, Charlson Comorbidity Index, Marital Status, Immigrant Status, Employment Status, Household Income, and Neighborhood Material Deprivation
\* $p < 0.05$ ; \*\* $p < 0.01$ ; \*\*\* $p < 0.0001$

**Supplementary Table 13:**
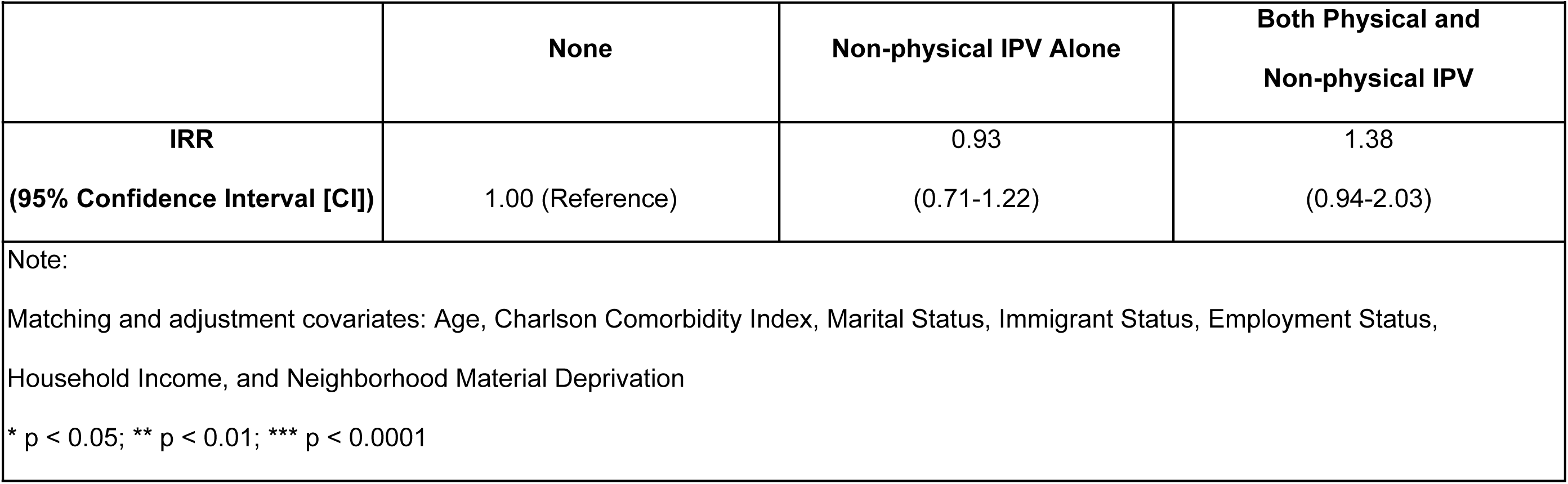
Adjusted Negative Binomial Regression Models for Cumulative Interactions with Ambulatory Care Over Study Period (Exposure to IPV within 2-years prior to participating in NEHW)

|  | None | Non-physical IPV Alone | Both Physical and<br>Non-physical IPV |
| --- | --- | --- | --- |
| **IRR**<br>**(95% Confidence Interval [CI])** | 1.00 (Reference) | 0.93<br>(0.71-1.22) | 1.38<br>(0.94-2.03) |

Note:
Matching and adjustment covariates: Age, Charlson Comorbidity Index, Marital Status, Immigrant Status, Employment Status, Household Income, and Neighborhood Material Deprivation
\* $p < 0.05$ ; \*\* $p < 0.01$ ; \*\*\* $p < 0.0001$

## References

1. Attala, J. M., Hudson, W. W., McSweeney, M., & Features Submission, H. C. (1994). A Partial Validation of Two Short-Form Partner Abuse Scales. Women & Health, 21(2–3), 125–139. 10.1300/J013v21n02_08

2. Azar, K. M. J., Petersen, J. P., Shen, Z., Nasrallah, C., Pesa, J., LaMori, J., & Pressman, A. (2020). Serious Mental Illness and Health-Related Factors Associated with Regional Emergency Department Utilization. Population Health Management, 23(6), 430–437. 10.1089/pop.2019.0161

3. Bonomi, A. E., Anderson, M. L., Rivara, F. P., & Thompson, R. S. (2009). Health Care Utilization and Costs Associated with Physical and Nonphysical-Only Intimate Partner Violence. Health Services Research, 44(3), 1052–1067. 10.1111/j.1475-6773.2009.00955.x

4. Bonomi, A. E., Trabert, B., Anderson, M. L., Kernic, M. A., & Holt, V. L. (2014). Intimate Partner Violence and Neighborhood Income: A Longitudinal Analysis. Violence Against Women, 20(1), 42–58. 10.1177/1077801213520580

5. Boyda, D., McFeeters, D., & Shevlin, M. (2015). Intimate partner violence, sexual abuse, and the mediating role of loneliness on psychosis. Psychosis, 7(1), 1–13. 10.1080/17522439.2014.917433

6. Canadian Institute for Health Information. (n.d.). National Ambulatory Care Reporting System (NACRS) metadata | CIHI. Retrieved June 11, 2024, from https://www.cihi.ca/en/national-ambulatory-care-reporting-system-nacrs-metadata

7. Canadian Institute for Health Information. (2024, February 22). NACRS emergency department visits and lengths of stay. https://www.cihi.ca/en/nacrs-emergency-department-visits-and-lengths-of-stay

8. Charlson, M., Carrozzino, D., Guidi, J., & Patierno, C. (2022). Charlson Comorbidity Index: A Critical Review of Clinimetric Properties. Psychotherapy and Psychosomatics, 91(1), 8–35. 10.1159/000521288

9. Coker, A. L. (2000). Physical Health Consequences of Physical and Psychological Intimate Partner Violence. Archives of Family Medicine, 9(5), 451–457. 10.1001/archfami.9.5.451

10. Cotter, A. (2021). Intimate partner violence in Canada, 2018: An overview. Juristat: Canadian Centre for Justice Statistics, 1,3–23.

11. Cunradi, C. B., Caetano, R., Clark, C., & Schafer, J. (2000). Neighborhood Poverty as a Predictor of Intimate Partner Violence Among White, Black, and Hispanic Couples in the United States: A Multilevel Analysis. Annals of Epidemiology, 10(5), 297–308. 10.1016/S1047-2797(00)00052-1

12. Dias, N. G., Costa, D., Soares, J., Hatzidimitriadou, E., Ioannidi-Kapolou, E., Lindert, J., Sundin, Ö., Toth, O., Barros, H., & Fraga, S. (2019). Social support and the intimate partner violence victimization among adults from six European countries. Family Practice, 36(2), 117–124. 10.1093/fampra/cmy042

13. Dichter, M. E., Sorrentino, A. E., Haywood, T. N., Bellamy, S. L., Medvedeva, E., Roberts, C. B., & Iverson, K. M. (2018). Women’s Healthcare Utilization Following Routine Screening for Past-Year Intimate Partner Violence in the Veterans Health Administration. Journal of General Internal Medicine, 33(6), 936–941. 10.1007/s11606-018-4321-1

14. Dusing, G. J., Essue, B. M., O’Campo, P., & Metheny, N. (2025). Long-term Public Healthcare Burden Associated with Intimate Partner Violence among Canadian Women: A Cohort Study. Health Policy, 105282. 10.1016/j.healthpol.2025.105282

15. Fedovskiy, K., Higgins, S., & Paranjape, A. (2008). Intimate Partner Violence: How Does it Impact Major Depressive Disorder and Post Traumatic Stress Disorder among Immigrant Latinas? Journal of Immigrant and Minority Health, 10(1), 45–51. 10.1007/s10903-007-9049-7

16. Freedman, D. A. (2006). On The So-Called “Huber Sandwich Estimator” and “Robust Standard Errors.” The American Statistician, 60(4), 299–302. 10.1198/000313006X152207

17. Goodman, L. A., & Epstein, D. (2022). Loneliness and the COVID-19 Pandemic: Implications for Intimate Partner Violence Survivors. Journal of Family Violence, 37(5), 767–774. 10.1007/s10896-020-00215-8

18. Gosangi, B., Lebovic, J., Park, H., Thomas, R., Gujrathi, R., Harris, M., Tornetta, P., & Khurana, B. (2021). Imaging patterns of lower extremity injuries in victims of intimate partner violence (IPV). Emergency Radiology, 28(4), 751–759. 10.1007/s10140-021-01914-5

19. Gujrathi, R., Tang, A., Thomas, R., Park, H., Gosangi, B., Stoklosa, H. M., Lewis-O’Connor, A., Seltzer, S. E., Boland, G. W., Rexrode, K. M., Orgill, D. P., & Khurana, B. (2022). Facial injury patterns in victims of intimate partner violence. Emergency Radiology, 29(4), 697–707. 10.1007/s10140-022-02052-2

20. Guttmann, A., Schull, M. J., Vermeulen, M. J., & Stukel, T. A. (2011). Association between waiting times and short term mortality and hospital admission after departure from emergency department: Population based cohort study from Ontario, Canada. BMJ (Clinical Research Ed.), 342, d2983. 10.1136/bmj.d2983

21. Hisasue, T., Kruse, M., Hietamäki, J., Raitanen, J., Martikainen, V., Pirkola, S., & Rissanen, P. (2024). Health-Related Costs of Intimate Partner Violence: Using Linked Police and Health Registers. Journal of Interpersonal Violence, 39(7–8), 1596–1622. 10.1177/08862605231211932

22. Iverson, K. M., Dardis, C. M., & Pogoda, T. K. (2017). Traumatic brain injury and PTSD symptoms as a consequence of intimate partner violence. Comprehensive Psychiatry, 74, 80–87. 10.1016/j.comppsych.2017.01.007

23. Izzy, S., Grashow, R., Radmanesh, F., Chen, P., Taylor, H., Formisano, R., Wilson, F., Wasfy, M., Baggish, A., & Zafonte, R. (2023). Long-term risk of cardiovascular disease after traumatic brain injury: Screening and prevention. The Lancet Neurology, 22(10), 959–970. 10.1016/S1474-4422(23)00241-7

24. Kanougiya, S., Sivakami, M., Daruwalla, N., & Osrin, D. (2022). Prevalence, pattern, and predictors of formal help-seeking for intimate partner violence against women: Findings from India’s cross-sectional National Family Health Surveys-3 (2005–2006) and 4 (2015–2016). BMC Public Health, 22(1), 2386. 10.1186/s12889-022-14650-3

26. Kirst, M., Lazgare, L. P., Zhang, Y. J., & O’Campo, P. (2015). The Effects of Social Capital and Neighborhood Characteristics on Intimate Partner Violence: A Consideration of Social Resources and Risks. American Journal of Community Psychology, 55(3), 314–325. 10.1007/s10464-015-9716-0

27. Leight, J., & Wilson, N. (2021). Intimate partner violence and maternal health services utilization: Evidence from 36 National Household Surveys. BMC Public Health, 21(1), 405. 10.1186/s12889-021-10447-y

28. Makaroun, L. K., Brignone, E., Rosland, A.-M., & Dichter, M. E. (2020). Association of Health Conditions and Health Service Utilization With Intimate Partner Violence Identified via Routine Screening Among Middle-Aged and Older Women. JAMA Network Open, 3(4), e203138. 10.1001/jamanetworkopen.2020.3138

29. Matheson, F. I., Dunn, J. R., Smith, K. L. W., Moineddin, R., & Glazier, R. H. (2012). Development of the Canadian Marginalization Index: A New Tool for the Study of Inequality. Canadian Journal of Public Health / Revue Canadienne de Sante’e Publique, 103, S12–S16.

30. Metheny, N., Dusing, G. J., Essue, B. M., & O’Campo, P. (2025). Non-Physical Intimate Partner Violence and Long-Term Public Healthcare Costs in a Representative Sample of Canadian Women. Violence Against Women. 10.1177/10778012251362231

31. O׳Campo, P., Wheaton, B., Nisenbaum, R., Glazier, R. H., Dunn, J. R., & Chambers, C. (2015). The Neighbourhood Effects on Health and Well-being (NEHW) study. Health & Place, 31, 65–74. 10.1016/j.healthplace.2014.11.001

32. Ohle, R., Ohle, M., & Perry, J. J. (2017). Factors associated with choosing the emergency department as the primary access point to health care: A Canadian population cross-sectional study. CJEM, 19(04), 271–276. 10.1017/cem.2016.350

33. Olive, P. (2018). Intimate partner violence and clinical coding: Issues with the use of the International Classification of Disease (ICD-10) in England. Journal of Health Services Research & Policy, 23(4), 212–221. 10.1177/1355819618781413

34. Peterson, A., Charles, V., Yeung, D., & Coyle, K. (2021). The Health Equity Framework: A Science– and Justice-Based Model for Public Health Researchers and Practitioners. Health Promotion Practice, 22(6), 741–746. 10.1177/1524839920950730

35. Pico-Alfonso, M. A., Garcia-Linares, M. I., Celda-Navarro, N., Blasco-Ros, C., Echeburúa, E., & Martinez, M. (2006). The Impact of Physical, Psychological, and Sexual Intimate Male Partner Violence on Women’s Mental Health: Depressive Symptoms, Posttraumatic Stress Disorder, State Anxiety, and Suicide. Journal of Women’s Health, 15(5), 599–611. 10.1089/jwh.2006.15.599

36. Rahman, B., Costa, A. P., Gayowsky, A., Rahim, A., Kiran, T., Ivers, N., Price, D., Jones, A., & Lapointe-Shaw, L. (2023). The association between patients’ timely access to their usual primary care physician and use of walk-in clinics in Ontario, Canada: A cross-sectional study. CMAJ Open, 11(5), E847–E858. 10.9778/cmajo.20220231

37. Saint-Eloi Cadely, H., Pittman, J. F., Pettit, G. S., Lansford, J. E., Bates, J. E., Dodge, K. A., & Holtzworth-Munroe, A. (2020). Temporal Associations Between Psychological and Physical Intimate Partner Violence: A Cross-Lag Analysis. Partner Abuse, 11(1), 22–38. 10.1891/1946-6560.11.1.22

38. Schull, M., Azimaee, M., Marra, M., Cartagena, R., Vermeulen, M., Ho, M., & Guttmann, A. (2019). ICES: Data, Discovery, Better Health. International Journal of Population Data Science, 4(2), 1135. 10.23889/ijpds.v4i2.1135

39. Sherin, K. M., Sinacore, J. M., Li, X. Q., Zitter, R. E., & Shakil, A. (1998). HITS: A short domestic violence screening tool for use in a family practice setting. Family Medicine, 30(7), 508–512.

40. Spencer, C. N., Khalil, M., Herbert, M., Aravkin, A. Y., Arrieta, A., Baeza, M. J., Bustreo, F., Cagney, J., Calderon-Anyosa, R. J. C., Carr, S., Chandan, J. K., Coll, C. V. N., de Andrade, F. M. D., de Andrade, G. N., Debure, A. N., Flor, L. S., Hammond, B., Hay, S. I., Knaul, F. N., … Gakidou, E. (2023). Health effects associated with exposure to intimate partner violence against women and childhood sexual abuse: A Burden of Proof study. Nature Medicine, 29(12), 3243–3258. 10.1038/s41591-023-02629-5

41. Stark, E. (2007). Coercive control: How men entrap women in personal life (pp. xii, 452). Oxford University Press.

42. Statistics Canada. (2019). 2011 National Household Survey (Nos. 99-014-X2011047).

43. Stubbs, A., & Szoeke, C. (2022). The Effect of Intimate Partner Violence on the Physical Health and Health-Related Behaviors of Women: A Systematic Review of the Literature. Trauma, Violence, & Abuse, 23(4), 1157–1172. 10.1177/1524838020985541

44. Sundararajan, V., Henderson, T., Perry, C., Muggivan, A., Quan, H., & Ghali, W. A. (2004). New ICD-10 version of the Charlson comorbidity index predicted in-hospital mortality. Journal of Clinical Epidemiology, 57(12), 1288–1294. 10.1016/j.jclinepi.2004.03.012

45. Valera, E. M., Campbell, J., Gill, J., & Iverson, K. M. (2019). Correlates of Brain Injuries in Women Subjected to Intimate Partner Violence: Identifying the Dangers and Raising Awareness. Journal of Aggression, Maltreatment & Trauma, 28(6), 695–713. 10.1080/10926771.2019.1581864

46. William, J., Loong, B., Hanna, D., Parkinson, B., & Loxton, D. (2022). Lifetime health costs of intimate partner violence: A prospective longitudinal cohort study with linked data for out-of-hospital and pharmaceutical costs. Economic Modelling, 116, 106013. 10.1016/j.econmod.2022.106013

47. World Health Organization. (2005). WHO multi-country study on women’s health and domestic violence against women: Summary report of initial results on prevalence, health outcomes and women’s responses. World Health Organization. https://iris.who.int/bitstream/handle/10665/43310/9241593512_eng.pdf?sequence=1

48. World Health Organization. (2013a). Global and regional estimates of violence against women: Prevalence and health effects of intimate partner violence and non-partner sexual violence. World Health Organization. https://iris.who.int/handle/10665/85239

49. World Health Organization. (2013b). Responding to intimate partner violence and sexual violence against women – Summary. https://www.who.int/publications-detail-redirect/WHO-RHR-13.10

50. World Health Organization. (2021). Violence against women prevalence estimates, 2018: Global, regional and national prevalence estimates for intimate partner violence against women and global and regional prevalence estimates for non-partner sexual violence against women. World Health Organization.

51. Wright, E. N., Anderson, J., Phillips, K., & Miyamoto, S. (2022). Help-Seeking and Barriers to Care in Intimate Partner Sexual Violence: A Systematic Review. Trauma, Violence, & Abuse, 23(5), 1510–1528. 10.1177/1524838021998305

52. Yakubovich, A. R., Heron, J., Metheny, N., Gesink, D., & O’Campo, P. (2021). Measuring the Burden of Intimate Partner Violence by Sex and Sexual Identity: Results From a Random Sample in Toronto, Canada –. Journal of Interpersonal Violence, 37(19–20). 10.1177/08862605211037433

53. Zhang, Y., Ancker, J. S., Hall, J., Khullar, D., Wu, Y., & Kaushal, R. (2020). Association Between Residential Neighborhood Social Conditions and Health Care Utilization and Costs. Medical Care, 58(7), 586. 10.1097/MLR.0000000000001337

